# The Mediating Role of Body Mass Index in the Association between Dietary Inflammatory Index and 10-year risk of CVD: a counterfactual mediation analysis

**DOI:** 10.1101/2023.06.06.23291060

**Authors:** Zechun Xie, Ling Wang, Mengzi Sun, Rui Wang, Yan Liu, Jing Li, Xuhan Wang, Ruirui Guo, Yuxiang Wang, Bo Li

## Abstract

**Introduction:** Dietary with higher inflammatory potential are associated with cardiovascular disease (CVD) risk over the next decade, while studies have shown body mass index (BMI) to be one of the mediators between dietary inflammatory index (DII) and multiple diseases such as depression and diabetes. However, the role of BMI in the association between DII and 10-year risk of CVD is unclear.

**Methods:** Study based on 14,355 adults from the National Health and Nutrition Examination Survey (NHANES). Participants’ diet, obesity level and CVD risk were assessed using the DII, BMI and Framingham Risk Score (FRS). Linear regression and counterfactual model were used to analyze this relationship.

**Results:** Participants on a pro-inflammatory diet had a higher risk of CVD over the next ten years compared to participants on an anti-inflammatory diet. Counterfactual models showed a 34.0% partial mediating role of BMI in this relationship, most particularly in males (17.3%) and non-elderly (28.6%-100%).

**Conclusion:** Inflammatory diet adversely affects cardiovascular risk, a large part of which operates through BMI, especially in males and non-elderly populations. These findings are beneficial in facilitating research on the mechanisms of diet-related inflammation on CVD.

## 1. Introduction

Cardiovascular disease (CVD) is a general term for diseases including coronary heart disease, cerebrovascular disease, hypertension, rheumatic heart disease and heart failure.[1] Nowadays, CVD poses a serious medical burden and is an important public health problem. Future deaths from CVD will continue to grow, with 17.5 million deaths from CVD globally in 2018[2], and are expected to increase to 23 million deaths by 2030[3]. CVD is caused by genetics and poor health behaviors or factors [4]. Previous findings reveal that modifiable lifestyles such as diet, smoking and obesity are strongly associated with cardiovascular disease or other disorders [5–7]. Khera et al. study found genetically similar two groups of people with poor health behaviors had nearly two times higher risk of coronary heart disease than those with ideal health behaviors [8].

Studies have shown that diets rich in saturated fatty acids, trans fatty acids, and salt are associated with a higher risk of CVD; conversely, an adequate intake of vegetables and fruits has a preventive effect on CVD [9, 10].As a modifiable lifestyle, increasing evidence also recognizes that pro-inflammatory foods enter the gut can induce inflammation and autoimmune responses, thereby accelerating the development of cardiovascular disease [11, 12]. Studies have shown that the effect of diet on cardiovascular disease is reflected in the result of the interaction of multiple food components rather than a single nutrient [13]. In addition, although several healthy dietary patterns, including the Mediterranean diet, have been shown to be associated with a reduced risk of CVD [14], the reduction in CVD risk by these healthy dietary patterns does not fully account for the role of anti-inflammatory foods. Therefore, we decided to use the dietary inflammation index developed by Shivappa et al. in our study to comprehensively assess the overall inflammatory potential of multiple dietary components.

A large intake of high-calorie and highly processed foods tends to cause obesity [9]. Visceral fat in obese individuals produces pro-inflammatory markers and is infiltrated by lymphocytes and macrophages, leading to inflammation. Clinical trials have also shown that modulating inflammation can prevent cardiovascular disease[15].Data from the Centers for Disease Control and Prevention point to a relative increase in heart disease and stroke mortality due to obesity in 2015, as well as a slower decline in coronary heart disease mortality and increased hospitalizations for heart failure in 2017, and these trends are also occurring among young adults [16, 17].

Several studies have also shown that body mass index (BMI) mediates the association between DII and a variety of diseases such as depression and diabetes [18, 19]. It is unclear whether BMI is an effect modifier in the association between DII and 10-year risk of CVD. Thus, we aimed to assess the association between DII and 10-year risk of CVD and to evaluate the interaction and mediating role of the mediating variable BMI in this association.

## 2. Methods

### 2.1 Study design and participants

Data for this study were obtained from NHANES in 2007-2016, with a total of 50,588 participants. The survey used complex probability sampling, and all publicly available data were collected by the National Center for Disease Control and Prevention through structured interviews and physical examinations[20]. The National Center for Health Statistics Institutional Ethics Board reviewed and approved the study protocol[21], and all participants provided written informed consent. Based on inclusion exclusion criteria, we excluded participants under 30 years of age, participants with cardiovascular disease (congestive heart failure, coronary artery disease, angina pectoris, heart disease and stroke), extreme dietary data (total energy intake <500 or >5000 kcal/day for females and <500 or >8000 kcal/day for males) and participants with missing information. Finally, 14,355 participants were included in this study (as shown in **Figure 1**).

**Figure 1.**
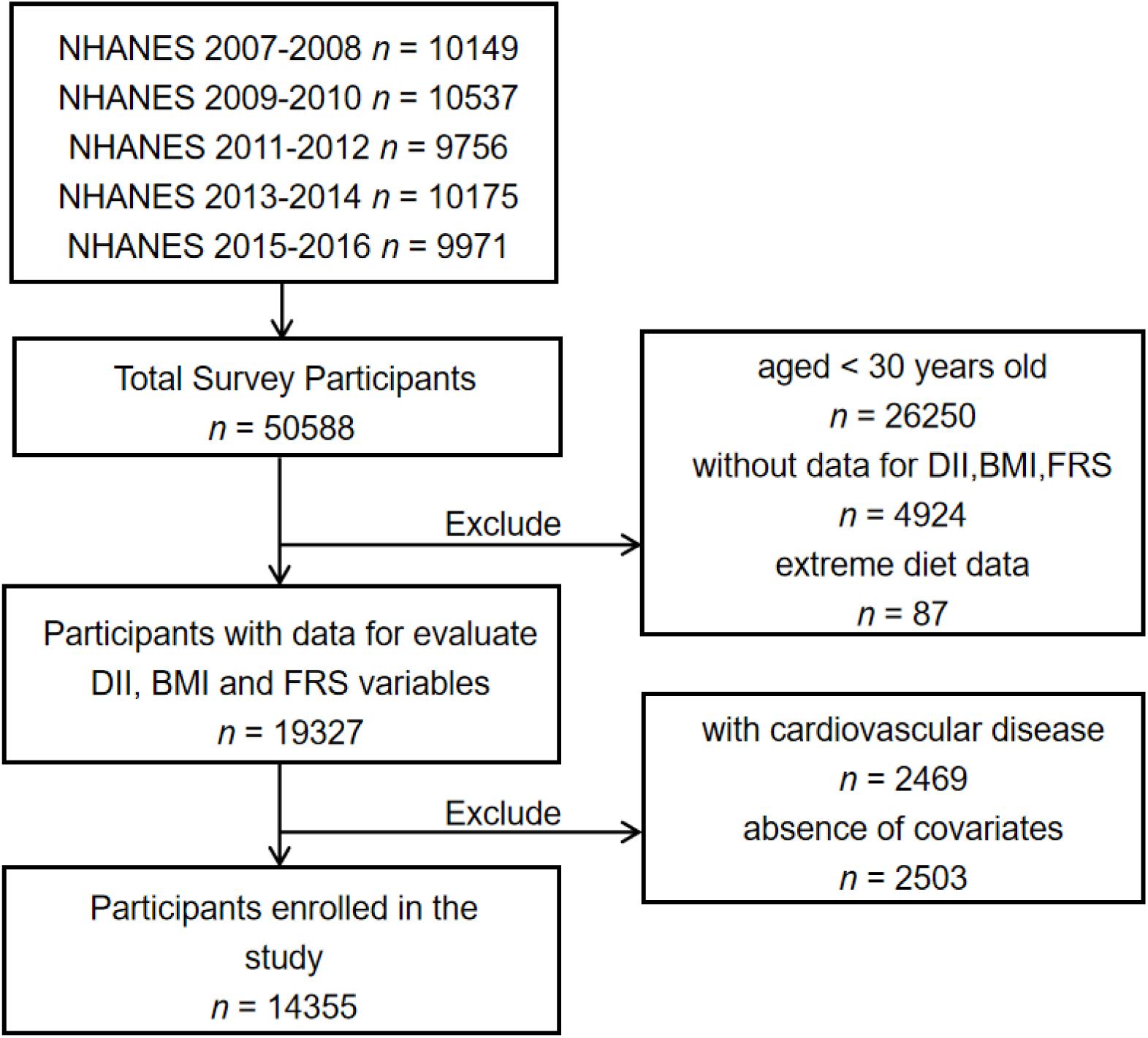
Flow chart of research subject selection. **Abbreviations:** DII, Dietary Inflammatory Index; BMI, body mass index; FRS, Framingham Risk Score.

### 2.2 Measurements

#### 2.2.1 Dietary Inflammatory Index (DII)

We used the dietary inflammatory index developed by Shivappa et al.[22] to assess the dietary inflammatory potential of individual participants. Dietary data were obtained via self-report, and dietary information for each participant was the average of two 24-hour dietary recall interviews. Dietary inflammatory levels were calculated using 45 food parameters that yielded participants’ exposure relative to the global standardized mean as a z-score. Z-scores were obtained by subtracting daily intake from a regionally representative database mean and dividing that value by the standard deviation of daily intake. The z-scores were then converted to a percentage (i.e., with values ranging from 0 to 1) and multiplied by 2 minus 1. This was done to minimize the effect of outliers or positive deviations. The resulting values were next multiplied by the corresponding food parameter scores and the scores of the 27 nutrients were summed to obtain the final values of the DII. The 27 nutrients used to calculate DII contained alcohol, vitamin B12/B6, β-carotene, caffeine, carbohydrate, cholesterol, total fat, fiber, folic acid, iron, magnesium, zinc, selenium, MUFA, niacin, n-3 fatty acids, n-6 fatty acids, protein, polyunsaturated fatty acids, riboflavin, saturated fat, thiamine, and vitamins A/C/D/E. The DII was calculated as DII ≥ 0 for a pro-inflammatory diet and DII < 0 for an anti-inflammatory diet[23].

#### 2.2.2 Body Mass Index

Height and weight were obtained by passing a standardized examination at a mobile examination center. BMI was calculated by dividing weight (kilogram) by the square of height (meters). In our analysis, BMI was included as a continuous variable.

#### 2.2.3 10-year risk of CVD

10-year risk of CVD was assessed using the Framingham Risk Score (FRS) [24]. The Framingham Risk Score is used to predict the risk of CVD over the next ten years in subjects over 30 years old who do not have cardiovascular disease. The algorithm uses the following measurements to predict 10-year risk of CVD: sex (male or female), age (years), HDL cholesterol (mg/dL), total cholesterol (mg/dL), diabetes status (yes or no, diabetes was defined as glycated hemoglobin ≥6.5%, or fasting glucose ≥126 mg/dL, or current insulin use), and smoking status (yes or no, smoking was defined as current smoker and nonsmoking was defined as former smoker and never smoker), treated or untreated (as determined by whether your doctor ever told you had hypertension and whether you were taking prescribed medications for hypertension) systolic blood pressure. The above parameters were measured by a health check-up. It is important to note that the total FRS scores for males and females correspond to different CVD risk, expressed as FRS%.

#### 2.2.4 Covariate assessment

We controlled for confounding factors identified in previous studies. Baseline characteristics included sex (male or female), age (30-44 years, 45-64 years, ≥65 years), education (<12 years, 12 years, ≥12 years), marital status (divorced, single, married), smoking status[25] (a person who has smoked 100 cigarettes in his life and now smokes was defined as a current smoker, a person who has smoked 100 cigarettes in his life but does not smoke now was defined as a former smoker, and a person who has not smoked 100 cigarettes in his life was defined as a non-smoker), drinking status [26](those who have had at least 12 alcoholic beverages in the past year are defined as current drinkers, those who have had at least 12 alcoholic beverages in their lifetime but not in the past year were defined as former drinkers, and those who have not had 12 alcoholic beverages in their lifetime were defined as non-drinkers), comorbidity[27] (classified as yes or no, and defined as hypertension, diabetes or hyperlipidemia. Hypertension were those who reported systolic blood pressure ≥ 140 mm Hg or diastolic blood pressure ≥ 90 mm Hg or taking anti-hypertensive medication. Diabetes were those who reported glycated hemoglobin (HbA1c) ≥ 6.5%, fasting blood glucose level ≥ 126 mg/dL, or current use of glucose-lowering medication. Hyperlipidemia were those who reported on lipid-lowering medications and total cholesterol concentration ≥ 240 mg/dL or a fasting LDL cholesterol concentration ≥ 160 mg/dL [28–30]), and energy intake (kcal).

### 2.3 Statistical analysis

In our analysis, continuous variables were calculated using means and standard errors and compared using t-tests for independent samples. Unweighted frequencies as well as weighted percentages and standard errors were calculated for categorical variables, and comparisons were made using the chi-square test. Linear regression models adjust for potential confounders were used to estimate the association of DII and BMI on 10-year risk of CVD and the interaction of DII and BMI. We analyzed the mediating role of BMI using a simple mediator model and a counterfactual model that allowed for exposure and mediator interactions. The mediation model includes three paths, paths a, b, and c. Path a is used to assess the relationship between DII (exposure) and BMI (mediation). Path b was used to assess the relationship between BMI (mediator) and 10-year risk of CVD (outcome). Path c (direct effect) assessed the effect of BMI on the association between DII and 10-year risk of CVD. The proportion of mediation was calculated as (pure natural indirect effect/total effect)*100%.

Statistical analyses were performed using the R package (http://www.R-project.org, The R Foundation) and SPSS 26.0. Two-tailed *P*-values <0.05 were considered statistically significant.

## 3. Results

### 3.1 Characteristics of Participants

Characteristics of participants with anti-inflammatory diet and pro-inflammatory diet are presented in **Table 1**. Of the 14355 participants, the overall mean age was 52.8 years, 48.8% were males, and the DII range was -5.11 to 5.17. In general, participants with pro-inflammatory diets were generally higher in BMI and 10-year risk of CVD, and dietary inflammatory potential was higher in females and younger participants. Additionally, the differences between anti-inflammatory and pro-inflammatory diets were statistically significant (*P* < 0.05) in terms of gender, age, energy intake, education level, marital status, smoking status, drinking status and comorbidity. Furthermore, there were also statistically significant differences in BMI and 10-year risk of CVD between anti-inflammatory and pro-inflammatory diets (*P* < 0.05).

**Table 1.**
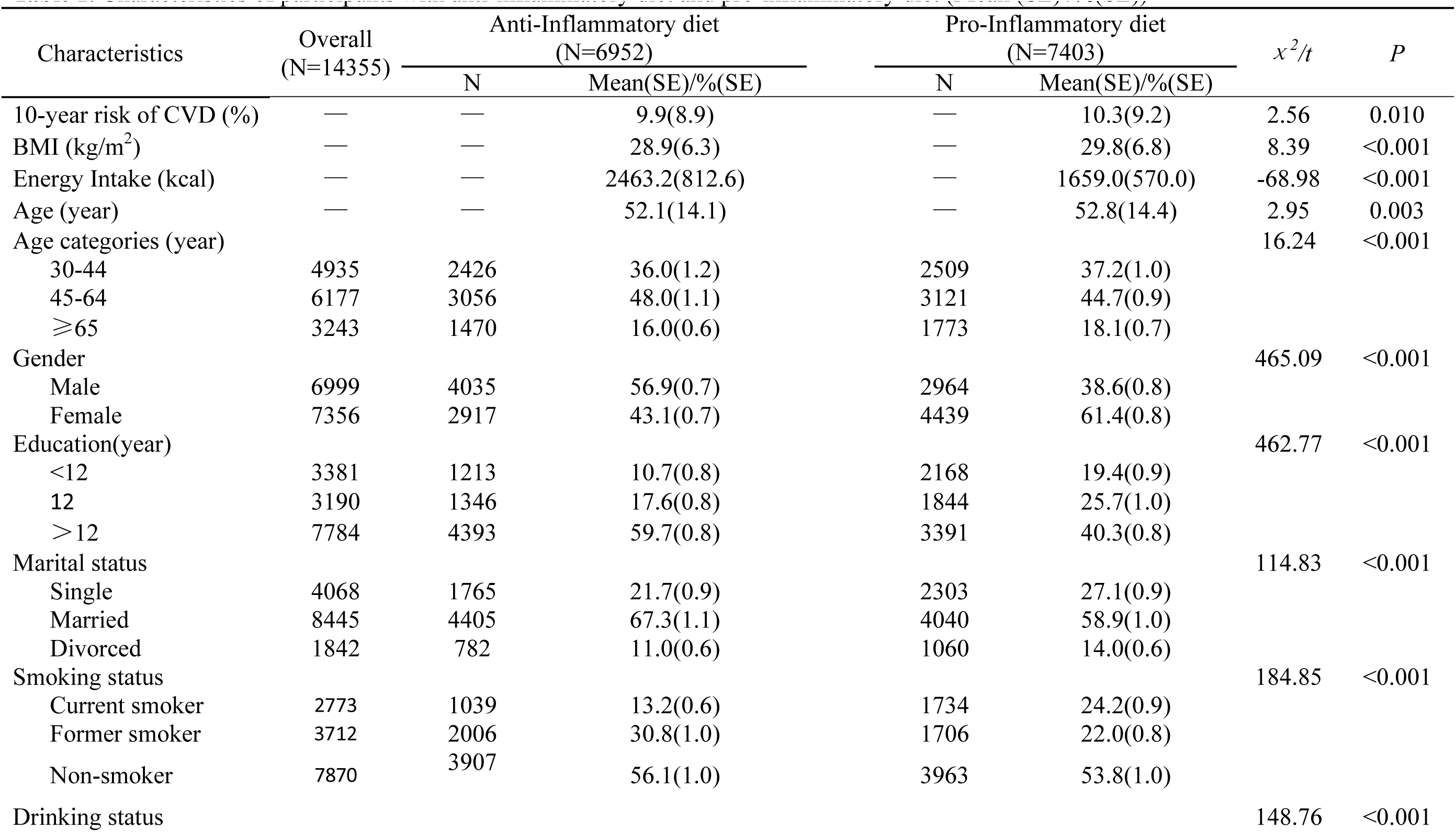
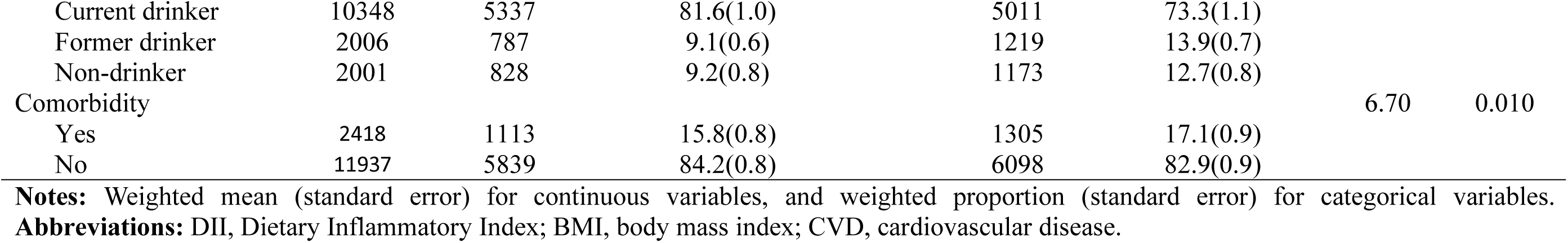
Characteristics of participants with anti-inflammatory diet and pro-inflammatory diet (Mean (SE) /%(SE))

The association between DII and BMI with 10-year risk of CVD is shown in **Table 2**. In the final model adjusted for confounders (sex, age, education level, marital status, smoking status, alcohol consumption, comorbidity and energy intake), DII and BMI were positively associated with 10-year risk of CVD *(P* < 0.01). In addition, there was an interaction between DII and BMI on 10-year risk of CVD (*P* = 0.005).

**Table 2.**
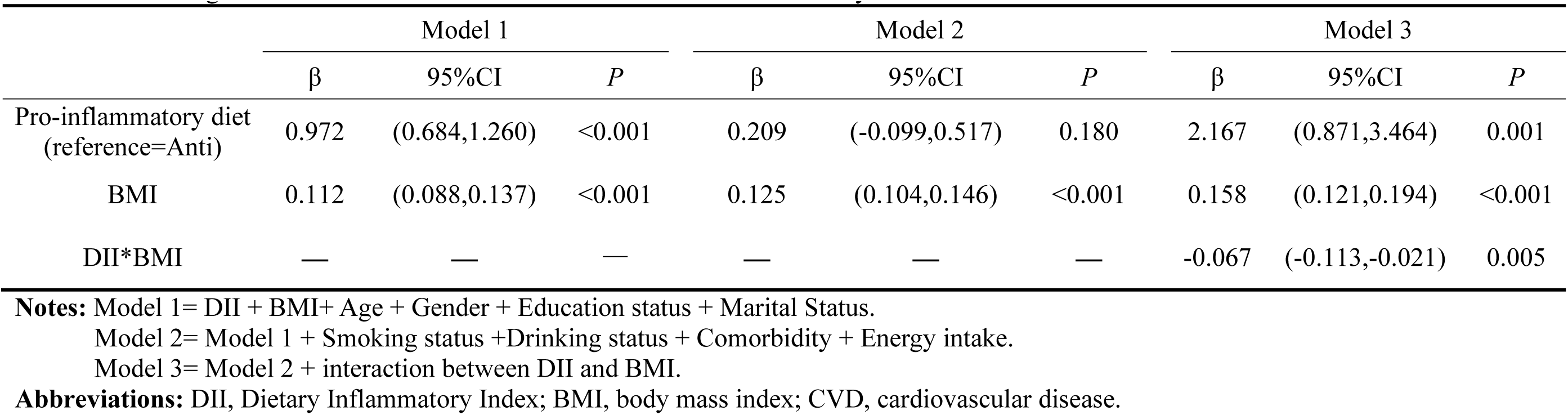
Linear regression models of associations between DII and BMI on 10-year risk of CVD.

### 3.2 Mediation of DII and 10-year risk of CVD Associations through BMI

We used BMI as a mediating variable to analyze the relationship between DII and 10-year risk of CVD. All mediation analyses were performed based on adjustment for sex, age, education level, marital status, smoking status, alcohol consumption, energy intake, and comorbidity. In the “traditional” mediation analysis model without the interaction of BMI and DII, increased DII was associated with increased 10-year risk of CVD, with 27.5% of the effect explained by a significant indirect effect of BMI (**Figure 2A**). Next, based on the existence of an interaction between DII and BMI, we used a counterfactual mediation model that allows for exposure-media interactions. In this model, BMI is not only a mediating variable but also a modifier of the effect of the relationship between DII and 10-year risk of CVD. We showed that in this model, a 34.0% association between DII and 10-year risk of CVD was mediated through BMI (**Figure 2B**).

**Figure 2.**
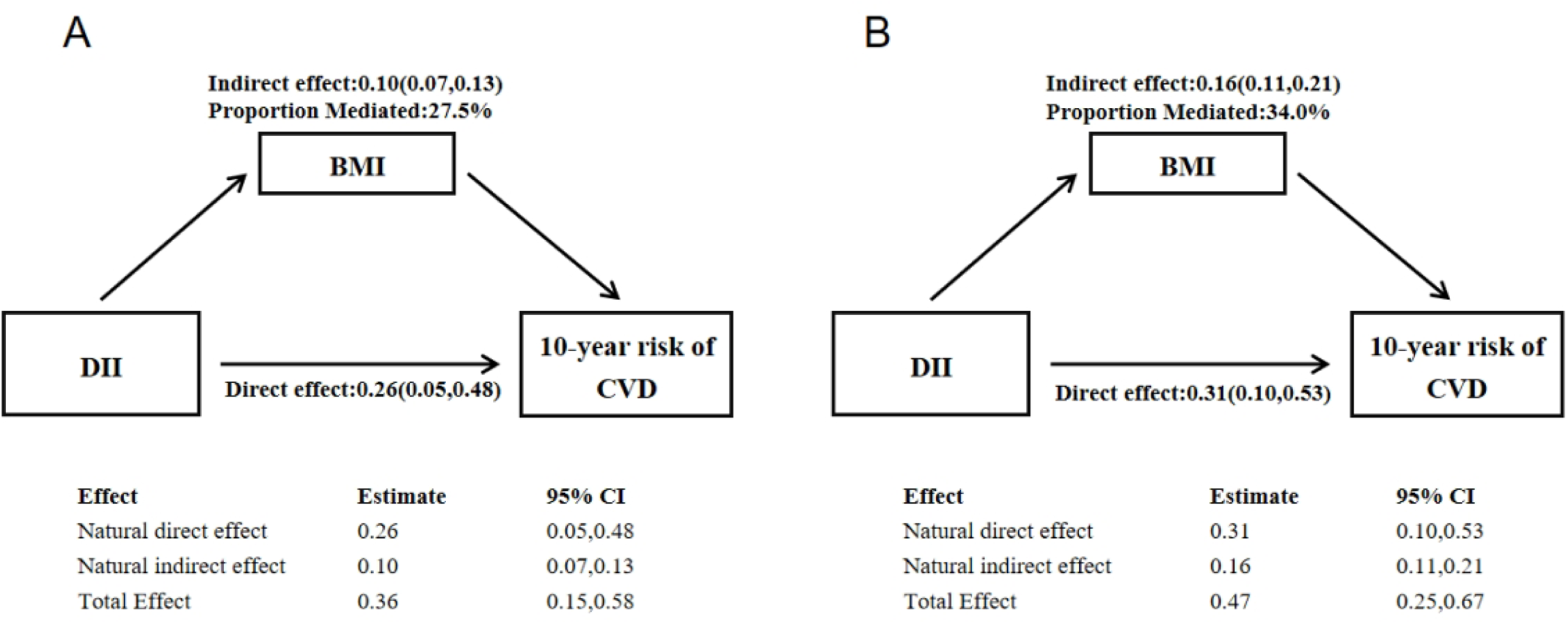
Mediation analyses without (A) and with (B) an interaction between DII and BMI on 10-year risk of CVD. **Notes:** Exposure: DII; Outcome: 10-year risk of CVD; Mediator: BMI. Model adjusted for sociodemographic variables (age, gender, education status, marital status), health behaviors (smoking status, drinking status), comorbidity, energy intake. **Abbreviations:** DII, Dietary Inflammatory Index; BMI, body mass index; CVD, cardiovascular disease; NDE natural direct effect, NIE natural indirect effect.

In a stratified analysis of the counterfactual mediation model, we found that 17.3% of the association between DII and 10-year risk of CVD was mediated by BMI in male participants, whereas we did not find any evidence of mediation of DII and 10-year risk of CVD by BMI in females. We also found that BMI played a partially mediating role in 28.6% of participants between the ages of 30-44 years, while it was fully mediating in participants between the ages of 45-64 years. We did not find evidence for an association of BMI mediating DII and 10-year risk of CVD in the older population (**Table 3**).

**Table 3.** Counterfactual mediation analysis of BMI on the association between DII and 10-year risk of CVD by Sex and Age Group Stratification.

|  | Sample<br>n | NDE | NIE | Total Effect | Proportion Mediated<br>(%) |
| --- | --- | --- | --- | --- | --- |
| **Stratification by Sex** |  |  |  |  |  |
| Males | 6999 | 0.62(0.30,0.95)*** | 0.13(0.07,0.21)*** | 0.75(0.41,1.09)*** | 17.3 |
| Females | 7356 | 0.11(-0.17,0.36) | 0.14(0.08,0.21)*** | 0.24(-0.03,0.52) | - |
| **Stratification by Age group** |  |  |  |  |  |
| 30-44 | 4935 | 0.35(0.20,0.51)*** | 0.14(0.08,0.21)*** | 0.49(0.31,0.66)*** | 28.6 |
| 45-64 | 6177 | 0.17(-0.20,0.51) | 0.25(0.16,0.37)*** | 0.42(0.01,0.79)* | 100 |
| >=65 | 3243 | 0.52(-0.04,1.06) | 0.06(0.00,0.17) | 0.60(0.03,1.13)* | - |

**Notes:** Exposure: DII; Outcome: 10-year risk of CVD; Mediator: BMI. Model adjusted for sociodemographic variables (age, gender, education status, marital status), health behaviors (smoking status, drinking status), comorbidity, energy intake. \*p < 0.05; \*\*p < 0.01; \*\*\*p < 0.001.
**Abbreviations:** DII, Dietary Inflammatory Index; BMI, body mass index; CVD, cardiovascular disease; NDE natural direct effect, NIE natural indirect effect.

## 4. Discussion

We analyzed the relationship between DII, BMI and 10-year risk of CVD in this study, and the following key findings were as followed. Firstly, we observed a positive association between pro-inflammatory diet and 10-year risk of CVD, and an interaction between DII and BMI on 10-year risk of CVD. In addition, using traditional mediator and counterfactual mediator model analyses, we found that this association was partially mediated by BMI and its interaction with DII. The mediating role of BMI accounted for 34.0% of this association in counterfactual model. In particular, when stratified by sex and age, a significant mediating effect of BMI was observed in males and in participants younger than 64 years.

More inflammatory food intake is associated with higher CVD risk [31]. Diet plays a critical role in the development of CVD as a modifiable exposure factor and as the most difficult heart-healthy behavior to achieve [32]. Previous study found that cardiovascular morbidity was 38% higher in those using a pro-inflammatory diet than in those consuming an anti-inflammatory diet, suggesting that reducing the intake of pro-inflammatory foods is an important strategy to prevent cardiovascular disease.[33]. Higher DII scores are associated with higher circulating inflammatory biomarkers (e.g., IL-1 and TNF-α), and higher concentrations of inflammatory biomarkers also migrate into vascular tissue [34], while damage to endothelial cells during the inflammatory response of the immune system increases the expression of cell adhesion molecules, which exacerbates the development of CVD by causing damage to the blood vessels.

Intervention studies have found that higher adherence to a Mediterranean diet reduces inflammation levels in people with high BMI [35, 36]. In another large cohort analysis of a middle-aged and older population, those randomly assigned to the Mediterranean diet had a 30% reduction in cardiovascular mortality, and this result was particularly evident in the obese population [37]. Obesity is a chronic inflammatory state resulting from immune system activation. Lipids, oxidized LDL particles and free fatty acids activate the immune inflammatory response, increase cardiovascular vascular inflammation, promote vascular injury and accelerate the onset of cardiovascular disease [38]. At the same time, similar healthy diets such as the Mediterranean diet can influence metabolic health and improve endothelial function through the gut microbiota [36]. Studies based on animal experiments and humans have shown that when the BMI is normal (between 18.5 and 24.9), adipose tissue is in an environment rich in anti-inflammatory cytokines, and conversely during elevated BMI adipocytes trigger an innate immune response that shifts the microenvironment to a pro-inflammatory state, producing and releasing cytokines such as TNF, IFNγ, IL-1β and IL-6, and this inflammatory change makes the body more susceptible to CVD[39].

However, more studies have shown heterogeneity in the association between BMI and CVD, being observed only in younger populations or in males [40, 41]. Interestingly, in our subgroup analysis, we had similar findings where obesity partially mediated 17.3%-28.6% of the association between DII and 10-year CVD risk in males and younger adults (30-44 years). This might be because men have a predominantly “apple” shape with more visceral adipose tissue, whereas women have a predominantly “pear-shaped obesity” with subcutaneous adipose tissue. The higher concentration of visceral adipose tissue ultimately leads to higher levels of free fatty acids and triglycerides in men than in women, which is associated with higher CVD[42]. The association between DII and CVD is partially mediated by BMI probably due to increased visceral fat in obese youth compared to their healthy weight peers[43]. And adult study has shown that excess abdominal visceral fat has the greatest adverse effect on CVD risk[44]. This finding suggests the importance of BMI control and pro-inflammatory diets in reducing cardiovascular disease risk in males and participants younger than 64 years. In addition, we found no direct association between DII and 10-year CVD risk in people aged 45-64 years, and the fully mediated role of BMI between DII and 10-year cardiovascular may provide new ideas for CVD prevention.

In the present study we found an interaction between dietary inflammation and BMI, and we suggest combining anti-inflammatory diet with maintaining a healthier weight to reduce CVD risk over the next decade. As the second most modifiable risk factor [45], it’s important to maintain a healthy weight to reduce the risk of CVD caused by a pro-inflammatory diet. Studies have shown that various types of weight loss interventions such as bariatric surgery, dietary interventions and pharmacological weight loss programs have shown beneficial effects on reducing cardiovascular risk [46–48]. Our study found that BMI significantly mediated the relationship between DII and 10-year risk of CVD. These results remind us that the harmful effects of pro-inflammatory diet and poor diet on CVD can be improved to some extent by controlling BMI.

Our study has several major strengths. First, our study sample was drawn from nationally representative decadal survey data and underwent rigorous and complex data collection, probability sampling, and review to reduce the effects of bias. Second, we used a dietary inflammation index to assess the overall inflammatory potential of foods rather than single nutrients, providing a more favorable persuasive approach to validating the diet-disease association. In addition, to our knowledge, this is the first exploration of the role of body mass index in the association between DII and cardiovascular risk in US adults over the next decade. However, our study also has limitations. First, dietary inflammatory indices were estimated by a 24-hour recall food questionnaire, which may have had some recall bias. Second, this was a cross-sectional study and more caution is needed when extrapolating the results. In conclusion, future prospective studies are necessary to overcome these limitations so that the results obtained can be repeatedly validated and extrapolated.

## 5. Conclusions

We elucidate that diets with high pro-inflammatory potential are associated with higher cardiovascular disease risk over the next decade, and that BMI may be one of the main mediators of this association, especially in men and non-elderly groups. Furthermore, our findings support current diet-based approaches to cardiovascular disease prevention. Our findings contribute to further studies on the mechanisms of action of diet-related inflammation affecting future cardiovascular risk.

## Authors’ contributions

All authors made appropriate contributions to the article. ZX, MS and BL were involved in the design of the study; ZX, MS and LW analyzed the data; ZX and LW wrote the manuscript; MS, LW, RW, JL, YL, XW, YD, RG, and YW were responsible for revising the manuscript. All authors agreed on the final version of the manuscript.

## Ethics approval and consent to participate

The protocols of NHANES were approved by the institutional review board of the National Center for Health Statistics, CDC. Written informed consent was obtained from each participant before participation in this study. ID: NCHS IRB/ERB Protocol #2011-17.

## Consent for publication

Each author has consented to the publication of this study

## Availability of data and material

Data described in the manuscript, code book, and analytic code will be made publicly and freely available without restriction at [https://www.cdc.gov/nchs/nhanes.

## References

1. Taylor F, Huffman MD, Macedo AF, Moore TH, Burke M, Davey Smith G, Ward K, Ebrahim S: Statins for the primary prevention of cardiovascular disease. Cochrane Database Syst Rev 2013, 2013(1):Cd004816.

2. Mendis S, Davis S, 9 2014; One More Landmark Step in the Combat Against Stroke and Vascular Disease. Stroke 2015, 46(5):E121–E122.

3. Mathers CD, Loncar D: Projections of global mortality and burden of disease from 2002 to 2030. Plos Med 2006, 3(11).

4. Naghavi M, Wang HD, Lozano R, Davis A, Liang XF, Zhou MG, Vollset SE, Ozgoren AA, Abdalla S, Abd-Allah F et al: Global, regional, and national age-sex specific all-cause and cause-specific mortality for 240 causes of death, 1990-2013: a systematic analysis for the Global Burden of Disease Study 2013. Lancet 2015, 385(9963):117–171.

5. Huxley RR, Woodward M: Cigarette smoking as a risk factor for coronary heart disease in women compared with men: a systematic review and meta-analysis of prospective cohort studies. Lancet 2011, 378(9799):1297–1305.

6. Stampfer MJ, Hu FB, Manson JE, Rimm EB, Willett WC: Primary prevention of coronary heart disease in women through diet and lifestyle. New Engl J Med 2000, 343(1):16–22.

7. Chiuve SE, McCullough ML, Sacks FM, Rimm EB: Healthy lifestyle factors in the primary prevention of coronary heart disease among men - Benefits among users and nonusers of lipid-lowering and antihypertensive medications. Circulation 2006, 114(2):160–167.

8. Khera AV, Emdin CA, Drake I, Natarajan P, Bick AG, Cook NR, Chasman DI, Baber U, Mehran R, Rader DJ et al: Genetic Risk, Adherence to a Healthy Lifestyle, and Coronary Disease. New Engl J Med 2016, 375(24):2349–2358.

9. Balakumar P, Maung-U K, Jagadeesh G: Prevalence and prevention of cardiovascular disease and diabetes mellitus. Pharmacol Res 2016, 113:600–609.

10. Balogun KA, Cheema SK: Cardioprotective Role of Omega-3 Polyunsaturated Fatty Acids Through the Regulation of Lipid Metabolism. Pathophysiology and Pharmacotherapy of Cardiovascular Disease:563–588.

11. Kriszbacher I, Koppan M, Bodis J: Inflammation, atherosclerosis, and coronary artery disease. New Engl J Med 2005, 353(4):429–429.

12. Libby P: Inflammation and cardiovascular disease mechanisms. Am J Clin Nutr 2006, 83(2):456s–460s.

13. Alissa EM, Ferns GA: Dietary fruits and vegetables and cardiovascular diseases risk. Crit Rev Food Sci 2017, 57(9):1950–1962.

14. Martinez-Gonzalez MA, Gea A, Ruiz-Canela M: The Mediterranean Diet and Cardiovascular Health. Circ Res 2019, 124(5):779–798.

15. Ferrucci L, Fabbri E: Inflammageing: chronic inflammation in ageing, cardiovascular disease, and frailty. Nat Rev Cardiol 2018, 15(9):505–522.

16. Ford ES, Ajani UA, Croft JB, Critchley JA, Labarthe DR, Kottke TE, Giles WH, Capewell S: Explaining the decrease in US deaths from coronary disease, 1980-2000. New Engl J Med 2007, 356(23):2388–2398.

17. Ford ES, Capewell S: Coronary heart disease mortality among young adults in the US from 1980 through 2002. Journal of the American College of Cardiology 2007, 50(22):2128–2132.

18. Laouali N, Mancini FR, Hajji-Louati M, El Fatouhi D, Balkau B, Boutron-Ruault MC, Bonnet F, Fagherazzi G: Dietary inflammatory index and type 2 diabetes risk in a prospective cohort of 70,991 women followed for 20 years: the mediating role of BMI. Diabetologia 2019, 62(12):2222–2232.

19. Ma Y, Li R, Zhan W, Huang X, Zhang Z, Lv S, Wang J, Liang L, Jia X: Role of BMI in the Relationship Between Dietary Inflammatory Index and Depression: An Intermediary Analysis. Front Med (Lausanne) 2021, 8:748788.

20. Centers for Disease Control and Prevention National Health and Nutrition Examination Survey [https://www.cdc.gov/nchs/nhanes.htm]

21. Centers for Disease Control and Prevention NCHS research ethics review board (ERB) approval [https://www.cdc.gov/nchs/nhanes/irba98.htm.]

22. Shivappa N, Steck SE, Hurley TG, Hussey JR, Hebert JR: Designing and developing a literature-derived, population-based dietary inflammatory index. Public Health Nutrition 2014, 17(8):1689–1696.

23. Davis JA, Mohebbi M, Collier F, Loughman A, Staudacher H, Shivappa N, Hebert JR, Pasco JA, Jacka FN: The role of diet quality and dietary patterns in predicting muscle mass and function in men over a 15-year period. Osteoporosis International 2021, 32(11):2193–2203.

24. D’Agostino RB, Vasan RS, Pencina MJ, Wolf PA, Cobain M, Massaro JM, Kannel WB: General cardiovascular risk profile for use in primary care - The Framingham Heart Study. Circulation 2008, 117(6):743–753.

25. Eke PI, Dye BA, Wei L, Slade GD, Thornton-Evans GO, Borgnakke WS, Taylor GW, Page RC, Beck JD, Genco RJ: Update on Prevalence of Periodontitis in Adults in the United States: NHANES 2009 to 2012. J Periodontol 2015, 86(5):611–622.

26. Chang HJ, Lin KR, Lin MT, Chang JL: Associations Between Lifestyle Factors and Reduced Kidney Function in US Older Adults: NHANES 1999-2016. Int J Public Health 2021, 66.

27. Lee Y, Son JS, Eum YH, Kang OL: Association of Sedentary Time and Physical Activity with the 10-Year Risk of Cardiovascular Disease: Korea National Health and Nutrition Examination Survey 2014-2017. Korean J Fam Med 2020, 41(6):374–380.

28. Zhao GX, Li CY, Ford ES, Fulton JE, Carlson SA, Okoro CA, Wen XJ, Balluz LS: Leisure-time aerobic physical activity, muscle-strengthening activity and mortality risks among US adults: the NHANES linked mortality study. Brit J Sport Med 2014, 48(3):244–249.

29. Lau E, Neves JS, Ferreira-Magalhaes M, Carvalho D, Freitas P: Probiotic Ingestion, Obesity, and Metabolic-Related Disorders: Results from NHANES, 1999-2014. Nutrients 2019, 11(7).

30. Schindler DK, Lopez Mitnik GV, Soliván-Ortiz AM, Irwin SP, Boroumand S, Dye BA: Oral Health Status Among Adults With and Without Prior Active Duty Service in the U.S. Armed Forces, NHANES 2011-2014. Mil Med 2020.

31. Yang QH, Cogswell ME, Flanders WD, Hong YL, Zhang ZF, Loustalot F, Gillespie C, Merritt R, Hu FB: Trends in Cardiovascular Health Metrics and Associations With All-Cause and CVD Mortality Among US Adults. Jama-J Am Med Assoc 2012, 307(12):1273–1283.

32. Anand SS, Hawkes C, de Souza RJ, Mente A, Dehghan M, Nugent R, Zulyniak MA, Weis T, Bernstein AM, Krauss RM et al: Food Consumption and its Impact on Cardiovascular Disease: Importance of Solutions Focused on the Globalized Food System A Report From the Workshop Convened by the World Heart Federation. Journal of the American College of Cardiology 2015, 66(14):1590–1614.

33. Li J, Lee DH, Hu J, Tabung FK, Li YP, Bhupathiraju SN, Rimm EB, Rexrode KM, Manson JE, Willett WC et al: Dietary Inflammatory Potential and Risk of Cardiovascular Disease Among Men and Women in the US. Journal of the American College of Cardiology 2020, 76(19):2181–2193.

34. Gordillo-Bastidas D, Oceguera-Contreras E, Salazar-Montes A, Gonzalez-Cuevas J, Hernandez-Ortega LD, Armendariz-Borunda J: Nrf2 and Snail-1 in the prevention of experimental liver fibrosis by caffeine. World J Gastroentero 2013, 19(47):9020–9033.

35. Richard C, Couture P, Desroches S, Lamarche B: Effect of the Mediterranean Diet with and Without Weight Loss on Markers of Inflammation in Men with Metabolic Syndrome. Obesity 2013, 21(1):51–57.

36. Schwingshackl L, Hoffmann G: Mediterranean dietary pattern, inflammation and endothelial function: A systematic review and meta-analysis of intervention trials. Nutr Metab Cardiovas 2014, 24(9):929–939.

37. Estruch R, Ros E, Salas-Salvado J, Covas MI, Corella D, Aros F, Gomez-Gracia E, Ruiz-Gutierrez V, Fiol M, Lapetra J et al: Primary Prevention of Cardiovascular Disease with a Mediterranean Diet Supplemented with Extra-Virgin Olive Oil or Nuts. N Engl J Med 2018, 378(25):e34.

38. Zhao Y, Qie RR, Han MH, Huang SB, Wu XY, Zhang YY, Feng YF, Yang XJ, Li Y, Wu YY et al: Association of BMI with cardiovascular disease incidence and mortality in patients with type 2 diabetes mellitus: A systematic review and dose-response meta-analysis of cohort studies. Nutr Metab Cardiovas 2021, 31(7):1976–1984.

39. Quail DF, Dannenberg AJ: The obese adipose tissue microenvironment in cancer development and progression. Nat Rev Endocrinol 2019, 15(3):139–154.

40. Zheng RZ, Zhou D, Zhu YM: The long-term prognosis of cardiovascular disease and all-cause mortality for metabolically healthy obesity: a systematic review and meta-analysis. J Epidemiol Commun H 2016, 70(10):1024–1031.

41. Yatsuya H, Folsom AR, Yamagishi K, North KE, Brancati FL, Stevens J, Investigators AS: Race- and Sex-Specific Associations of Obesity Measures With Ischemic Stroke Incidence in the Atherosclerosis Risk in Communities (ARIC) Study. Stroke 2010, 41(3):417–425.

42. Rana MN, Neeland IJ: Adipose Tissue Inflammation and Cardiovascular Disease: An Update. Curr Diab Rep 2022, 22(1):27–37.

43. Higgins S, Zemel BS, Khoury PR, Urbina EM, Kindler JM: Visceral fat and arterial stiffness in youth with healthy weight, obesity, and type 2 diabetes. Pediatr Obes 2022, 17(4):e12865.

44. Cruickshank K, Riste L, Anderson SG, Wright JS, Dunn G, Gosling RG: Aortic pulse-wave velocity and its relationship to mortality in diabetes and glucose intolerance: an integrated index of vascular function? Circulation 2002, 106(16):2085–2090.

45. Lavie CJ, Arena R, Alpert MA, Milani RV, Ventura HO: Management of cardiovascular diseases in patients with obesity. Nat Rev Cardiol 2018, 15(1):45–56.

46. Aggarwal R, Harling L, Efthimiou E, Darzi A, Athanasiou T, Ashrafian H: The Effects of Bariatric Surgery on Cardiac Structure and Function: a Systematic Review of Cardiac Imaging Outcomes. Obes Surg 2016, 26(5):1030–1040.

47. Osei-Assibey G, Boachie C: Dietary interventions for weight loss and cardiovascular risk reduction in people of African ancestry (blacks): a systematic review. Public Health Nutrition 2012, 15(1):110–115.

48. Kane JA, Mehmood T, Munir I, Kamran H, Kariyanna PT, Zhyvotovska A, Yusupov D, Suleman UJ, Gustafson DR, McFarlane SI: Cardiovascular Risk Reduction Associated with Pharmacological Weight Loss: A Meta-Analysis. International journal of clinical research & trials 2019, 4(1).

